# Optimizing Contingency Management with Reinforcement Learning

**DOI:** 10.1101/2024.03.28.24305031

**Authors:** Young-geun Kim, Laura Brandt, Ken Cheung, Edward V. Nunes, John Roll, Sean X. Luo, Ying Liu

**Author notes:** **Corresponding Authors:** Sean X. Luo, M.D., Ph.D., 1051 Riverside Dr, New York, NY 10032, Ying Liu, Ph.D., 722 West 168th street, R214, New York, NY 10032.

## Abstract

Contingency Management (CM) is a psychological treatment that aims to change behavior with financial incentives. In substance use disorders (SUDs), deployment of CM has been enriched by longstanding discussions around the cost-effectiveness of prized-based and voucher-based approaches. In prize-based CM, participants earn draws to win prizes, including small incentives to reduce costs, and the number of draws escalates depending on the duration of maintenance of abstinence. In voucher-based CM, participants receive a predetermined voucher amount based on specific substance test results. While both types have enhanced treatment outcomes, there is room for improvement in their cost-effectiveness: the voucher-based system requires enduring financial investment; the prize-based system might sacrifice efficacy. Previous work in computational psychiatry of SUDs typically employs frameworks wherein participants make decisions to maximize their expected compensation. In contrast, we developed new frameworks that clinical decision-makers choose actions, CM structures, to reinforce the substance abstinence behavior of participants. We consider the choice of the voucher or prize to be a sequential decision, where there are two pivotal parameters: the prize probability for each draw and the escalation rule determining the number of draws. Recent advancements in Reinforcement Learning, more specifically, in off-policy evaluation, afforded techniques to estimate outcomes for different CM decision scenarios from observed clinical trial data. We searched CM schemas that maximized treatment outcomes with budget constraints. Using this framework, we analyzed data from the Clinical Trials Network to construct unbiased estimators on the effects of new CM schemas. Our results indicated that the optimal CM schema would be to strengthen reinforcement rapidly in the middle of the treatment course. Our estimated optimal CM policy improved treatment outcomes by 32% while maintaining costs. Our methods and results have broad applications in future clinical trial planning and translational investigations on the neurobiological basis of SUDs.

## INTRODUCTION

Substance use disorders (SUDs) are behavioral pathologies that involve common underlying neurobiological substrates involving the dopaminergic circuitry in the midbrain (1,2). This circuit has been hypothesized to allow the brain to learn from and process rewarding experiences (3– 5). A substantial body of literature has accumulated around interrogating animal models to elucidate the role of dopamine and prediction error in SUDs (6). However, translating these insights to clinical settings represents a substantial challenge, particularly because the meaning of these neural signatures has limited known behavioral correlates in the real world. With an increasing number of lives affected by SUD-related complications and overdoses (7), there is an urgency in developing novel SUD treatments (8). Contingency Management (CM), which uses financial incentives to promote substance abstinence, stands out as a promising approach (9– 15) with the potential to enhance public health through broad dissemination. Furthermore, CM represents a natural translational foundation to connect preclinical neuroscience to clinical applications.

There is ample evidence supporting CM’s effectiveness in treating a variety of SUDs, including alcohol (16), benzodiazepines (17), stimulants (18,19), cannabis (20,21), and opioid (22,23) use disorders. Its efficacy extends to diverse populations, including pregnant women (24,25) and adolescents (26). Common CM schemas can be classified into voucher-based (27– 29) and prize-based (16,30,31) approaches. Voucher-based methods reward participants with vouchers of escalating value to redeem goods or services for submitting substance-free biological samples (32). Prize-based approaches, designed to mitigate the high incremental costs associated with vouchers, let participants draw for prizes using a lottery based on their urine test results (33). Meta-analyses demonstrated that both approaches are effective in encouraging SUD remission (34–39).

Broad dissemination of CM treatment requires a balance between cost and efficacy. The high cost of voucher-based CM is a barrier to its widespread clinical adoption (40) and generates organizational (41) and legal challenges (42). A lower cost prized-based schema, however, might sacrifice efficacy (43): in general, higher financial reinforcement correlates to better outcomes (44–46). Clinical trials examine one or a few alternative treatment schemas at a time, with the schemas themselves determined by the clinicians and researchers ad hoc based on clinical feasibility and historical designs.

Computationally, voucher-based and prize-based systems represent two small regions in a high dimensional space that captures all possible CM schemas in a treatment process. No existing study has compared a treatment-as-usual schema with the theoretically optimal schema. This is due to the lack of a mathematically precise method to capture the schemas and make computational inferences, as well as practical limitations in conducting clinical studies to evaluate all possible CM schemas.

In response to this knowledge gap, we developed a new computational psychiatry framework using Reinforcement Learning to assess different classes of CM schemas using a CM clinical trial dataset. Reinforcement Learning is a powerful data science framework to analyze such dynamic treatment regimens in precision medicine (47), which typically focuses on identifying optimal rules to maximize treatment outcomes (48). While existing frameworks in computational psychiatry usually model participants’ decision-making to understand the mechanism of receiving compensation (5), our approach models the clinical treatment process to directly target cost-effectiveness and imposes budget limitations on CM schemas to identify optimal ones within these. We applied the off-policy evaluation techniques (49) to predict the outcomes of new CM schemas. We used a pragmatic clinical trial from the National Drug Treatment Clinical Trials Network (CTN-0007) trial (50) data, which contained detailed, week-by-week urine toxicology and prize-based rewards. We hypothesized that this method would allow us to identify the most cost-effective strategies that meet specified budget constraints.

## METHODS

### Participants and Study Design

The CTN-0007 trial employed a prize-based CM schema targeting stimulant abstinence in individuals undergoing methadone maintenance treatment. Table 1 summarized the trial structure and cohort characteristics. The CTN-0007 trial is a community-based methadone maintenance treatment with 388 participants from six clinics, enhancing the main treatment outcomes. Participants visited local clinics or treatment programs twice weekly, yielding 24 visits over 12 weeks. At each visit, they provided biological samples such as breathalyzer tests for alcohol and urine tests for opioids. Participants who achieved substance abstinence since their last visit earned draws and had the opportunity to receive motivational incentives based on a prize structure. The prizes included: (i) 50% of tickets with a “good job” message without tangible incentive ($0 value), (ii) 41.8% of tickets with small incentives ($1 value), (iii) 8% of tickets with large incentives ($20 value), and (iv) 0.2% of tickets with jumbo incentives ($80 value). The primary substance use test was considered negative if all primary substances (alcohol, amphetamine, cocaine, and methamphetamine) yielded negative results. The escalation of prizes was linear: the number of draws increased by one as the abstinence period increased by one week, and it was reset to zero if the primary substance test result was positive. Two bonus draws were provided if test results for all substances were negative. Fully anonymized CTN-0007 data were obtained through the publicly accessible CTN data repository: http://datashare.nida.nih.gov/study/nida-ctn-0007.

### Reinforcement Learning Framework for CM Schemas

The CTN-0007 CM schema, which was applied to several other studies (21,33), sequentially offered incentives to reinforce substance abstinence based on prior substance testing records. This CM schema was, therefore, a dynamic treatment program (51–53), a sequence of decision rules tailored to individuals’ time-varying substance use history at each visit time.

We formulated elements in CM from a Reinforcement Learning perspective: The CM schema was converted into a set of decision rules (corresponding to “policy” in Reinforcement Learning terminology (54)) that took the individuals’ substance use histories (“state”) at each visit and mapped them to the incentives (“actions”). CM schema varied based on the probabilities of winning each prize and the escalation rule for the number of draws according to substance use histories. Our framework modeled voucher-based approaches by setting the probability of getting a specific prize to 1. Substance use test results at the next visit were either positive or negative (corresponding to “reward” in the Reinforcement Learning framework, not to be confused with reward in financial incentives). The efficacy of CM during the whole treatment program was evaluated by summing the total number of negative substance test samples (“discounted cumulative reward” (54)). We summarized our framework in Figure 1.

We provided basic notations with assumptions. Our framework denoted the incentive and substance use history at the *t*-th visit by *A*_*t*_ asnd *S*_*t*_, respectively. We assumed that *A*_*t*_ and *S*_*t*_ had finite supports and 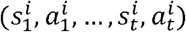 was independent and identically distributed, where *i* denoted the patient index. The CM schema applied at the *t*-th visit was denoted by 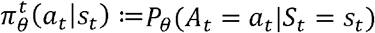 where 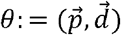 represented the time-invariant CM parameter and 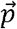 was the vector of prize probabilities. Additionally, 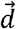 was the vector of the number of draws given the longest duration of substance abstinence. We described details on the CM parameterization in the next subsection. The reward was denoted by *R*_*t*_. We provided further details on our Reinforcement Learning framework in Appendix 1.

### Parameterization of CM Schemas

We parameterized a range of CM schemas 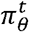 and analyzed the performance of these hypothetical CM schemas. CM schema parameters included (i) prize probabilities 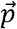 and (ii) the number of draws given the longest duration of substance abstinence 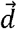. Reflecting the CTN-0007 study that had four prize types and was a twelve-week program, the 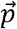 was a four-dimensional vector whose elements were non-negative and their sum was 1 and 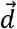 was a twelve-dimensional vector whose *k*-th element was the ticket number when the longest duration of substance abstinence was *k* weeks. With these parameters, the 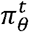 was expressed as follows:

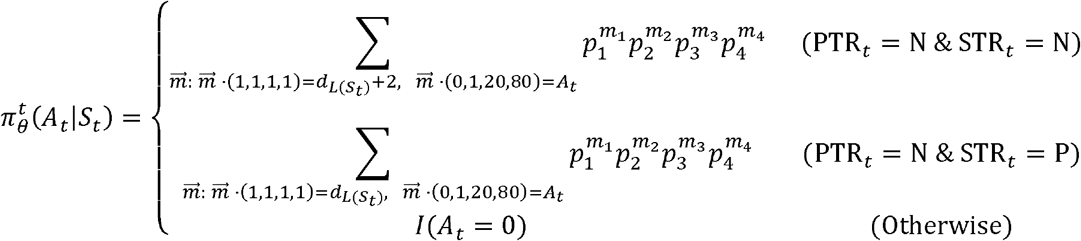

where *L*(*S*_*t*_)denoted the longest duration of primary substance abstinence and 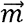 denoted the four-dimensional vector consisting of the number of drawn prizes. Additionally, the PTR_*t*_ and STR_*t*_ indicated the primary and secondary substance test results at the *t*-th visit, respectively, determined by *S*_*t*_. In this parameterization, the 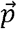 affected on the probability mass 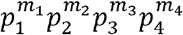 for given lottery events and the 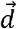 affected on the set of lottery events whose probability masses were summed up. For example, the set was 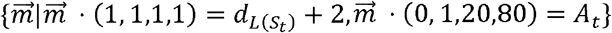 when both primary and secondary substance test results were negative, reflecting the 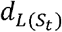 and two draws for primary and secondary substance abstinences, respectively.

### Evaluating New CM Schemas using Off-policy Evaluation

We used Off-Policy Evaluation, a set of techniques in Reinforcement Learning that estimates the effectiveness of new policies with data collected from existing fixed policies (49), to evaluate parameterized hypothetical CM schemas. The CM schema 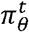 described incentive structures given substance use records and its performance was evaluated with the *value function* (54), the expected total discounted reward, defined as 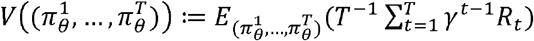

Here, *T* was the number of total visit, 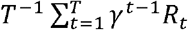 was the total discounted reward, and γ was the discount factor to balance rewards from early and late stages. Given that all the negative test results equally contributed to the number of substance-free samples, which is the same as the proportion of substance-free samples up to constant multiplication, we set the value of γ to 1.

We used the importance sampling estimator (55), an off-policy evaluation method, to evaluate hypothetical CM schemas with the CTN-0007 data. For a given target CM schema with parameter *θ*_*Tar*_ (e.g., previously unstudied prize probabilities), the importance sampling estimator for the value of the target schema 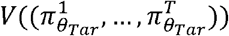 was unbiased and was expressed as follows:

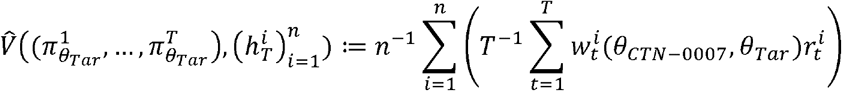

where 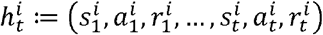 was collected under the existing schema 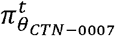, the CM schema used in CTN-0007, *θ*_*CTN*-0007_ was the parameter for the existing schema, *n* was the sample size, and 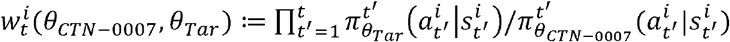 was the importance weight, representing the relative likelihood of data under the target CM schema compared to the existing schema. Note that both target and existing schemas were tractable, so no further model approximation was required to compute the importance weights.

### Optimizing CM Schema Under Cost Constraint

We imposed a specified budget and the set of CM parameters to optimize. We defined the estimated optimal CM parameter under the budget level *B* with data 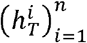 as follows:

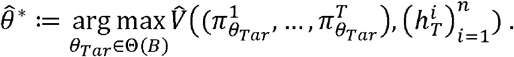

where Θ,(B), indicated the set of feasible parameters under the budget *B*. We first specified a grid for parameters, described in the next subsection, and then filtered the grid using bootstrap confidence intervals (CIs) of treatment costs. With 1,000 bootstrapped datasets from CTN-0007, we calculated 95% bootstrap CIs of cost for a particular CM schema and removed the schema if the upper bound was larger than the budget. Among CM parameters in Θ(*B*), we searched for the one that maximized value functions, the total number of primary substance-free samples (up to constant multiplication), estimated with the bootstrap mean for importance sampling estimators 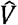 using incentive group in CTN-0007.

### Search Grid

We considered a sixteen-dimensional search grid for parameters *θ*; the first four were for prize probabilities 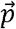 and the last twelve were for the number of draws given the longest duration of substance abstinence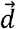. The search grid was illustrated in Figure 2(a). For the 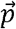, we considered 1,000 points at an intuitive curve connecting prize probabilities for usual care (UC) and for CM schema used in CTN-0007. For the 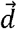, we considered three options for escalation of prizes: (i) linear escalation rule in CTN-0007, (ii) logarithmic escalation, and (iii) logistic escalation. The rules were parameterized to have total ticket numbers comparable to the linear rule. The linear escalation rule increased the number of draws at a constant speed. The logarithmic escalation rule rapidly increased the number of draws early in treatment and kept decreasing the speed of increment. The logistic escalation rule gradually increased prize draws early in treatment, rapidly increased them in the middle of treatment, and then gradually increased again to converge to the number of draws by the linear rule in the late treatment. We provided details on the search grid in Appendix 2.

## RESULTS

### Lower Prize Expectation Does Not Sacrifice Treatment Effect

We examined the marginal effect of prize probabilities. The comparison between CM schemas with diverse 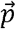 while fixing 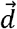 as the existing linear rule was shown in Figure 2(b). Hypothetical CM schemas with prize probabilities closer to the value for usual care (UC), 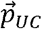 had lower prize expectations per each draw. The CM schema with the 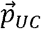 had significantly lower treatment costs (p<0.001^1^; mean difference=-$117.8; 95% CI=[-$149.13, -$91.80]) without notable changes on levels of treatment outcomes (p=0.119; mean difference=-1.61; 95% CI=[-4.08, 1.22]).

### Logistic Escalation Rule Is More Cost Effective Than Existing Linear Rule

We examined the marginal effect of escalation rules for the number of draws given the longest duration of substance abstinence. The comparison between CM schemas with diverse 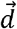 while fixing 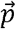 as the probability of the CM schema in CTN-0007 was shown in Figure 2(c). The hypothetical CM schema using the logistic escalation rule achieved superior treatment results (p<0.001; mean difference=3.72; 95% CI=[1.34, 6.15]) without a significant cost increase relative to the existing linear escalation rule (p=0.075; mean difference=$30.92; 95% CI=[-$12.15, $73.66]): the average number of primary substance-free samples under the logistic escalation rule increased by 43% compared to the existing linear escalation rule. However, the budget, the upper bound of 95% CIs of costs, was larger than that of the existing schema. Conversely, the logarithmic escalation rule exhibited similar treatment outcomes but incurred higher costs.

### Optimal Policies under Different Budget Constraints

We identified the optimal CM schema under the budget used in CTN-0007. The comparison between existing and optimal CM schemas was shown in Figure 2(d). The optimal schema achieved superior treatment results with comparable costs relative to the existing schema while satisfying the budget constraint. Detailed numerics were provided in Table 2. Table 2 described the comparative analysis results among three distinct CM schemas: (i) existing CM schema implemented in CTN-0007, (ii) the optimal CM constrained by the CTN-0007 budget ($149), and (iii) the optimal CM bounded by the annual cap established by Washington State ($100) (42).

Our results showed that schema (ii) had a significantly larger number of primary substance-free samples (p=0.007; mean difference=2.77; 95% CI=[0.53, 5.06]), increasing by 32%, without a significant increase in costs (p=0.419; mean difference=$3.74; 95% CI=[-$36.02, $42.12]) compared to (i). Notably, schema (iii), despite its tighter budget, outperformed schema (i) in terms of treatment costs (p=0.013; mean difference=-$37.38; 95% CI=[-$73.33, -$4.31]) without significant sacrifice in treatment outcomes (p=0.099; mean difference=1.38; 95% CI=[-0.70, 3.51]). With the same budget used in schema (i), schema (ii) and (iii) modified the incentive allocation by lowering the prize expectation and by shifting more lottery tickets from early on in treatment to later in treatment.

## DISCUSSION

In this study, we leveraged Reinforcement Learning techniques to assess CM schemas in treating SUDs and identified optimal CM schemas under budget constraints. We found that the CM schema using the logistic escalation rule was estimated to have a 43% improvement in negative urine drug screens and negative breathalyzer samples over a three-month study period compared to the existing CM schema in CTN-0007 using a linear program. Imposing a cost constraint, we showed that an optimal CM schema with logistic rule and the probability of getting $0, $1, $20, and $80 prizes of 55.40%, 37.28%, 7.14%, and 0.18%, respectively, was estimated to have a 32% improvement in total number of negative urine drug screens and negative breathalyzer over a three-month study period. Our findings on the marginal effect of prize probabilities align with established literature, illustrating that potent reinforcers boost treatment outcomes and raise costs. Moreover, our analysis highlighted the superior cost-effectiveness of the logistic escalation rule compared to its linear and logarithmic counterparts. The CM schemas we identified optimize cost-effectiveness, maximizing outcomes within budgetary constraints.

Considering more CM parameters may further enhance the efficacy of our approach. For example, we can add parameters for the maximum prize amount and the choice of primary and secondary substances for future research. We assumed that the patients were not sensitive to the subtle difference between the informed and actually applied CM schemas.

We can deliver the optimal CM schema identified by our approach in clinical settings. To validate our findings, conducting a Randomized Controlled Trial (RCT) is an essential future direction. Implementing previously unstudied CM schema, such as the logistic escalation rule, potentially enhances treatment efficacy. Additionally, our approach applies to broader clinical settings, allocating resources depending on the patient’s status. For example, the CTN-0013 study (56) provided Motivational Enhancement Therapy for pregnant substance users (MET-PS). We can extend our Reinforcement Learning framework and analyze the effectiveness of MET-PS by viewing the schema of providing MET-PS as a policy using participants’ substance use histories.

Our Reinforcement Learning framework also allows time-varying personalized CM schemas, promising to provide tailored treatment interventions. For example, participants who abstain from opioid use in the first three weeks were less likely to return to opioid use (57). We can apply two CM parameters: one is for these early stages, and the other is for the remaining stages to implement time-varying CM schemas. Another example is the CTN-0013 study, where the efficacy of MET-PS varied between sites and between majority and minority groups. We can cluster participants according to their demographic or economic characteristics and apply cluster-specific CM parameters to implement personalized CM schemas.

## Data Availability

All data produced are available online at http://datashare.nida.nih.gov/study/nida-ctn-0007.

## ACKNOWLEDGEMENT

Young-geun Kim and Ying Liu were supported by NIH R01MH124106.

## APPENDIX

### Appendix 1. Details on Reinforcement Framework for CM Schemas

Clinical trial data were encoded using our framework, illustrated in Table 3. An example of a patient’s trajectory under a CM schema was shown: on their 4^th^ visit, *A*_4_ = $21 was provided based on the lottery results whose number of draw was determined by *S*_4_. The “reward” *R*_4_ was 1 since the primary substance test result at the 5th visit was negative. This example suggested that the action *A*_4_ of $21 strengthened the primary substance abstinence, resulting in *R*_4_ of 1. In contrast, in the 2nd and 3rd visits, incentives of $2 and $1, respectively, were enough to maintain primary substance abstinence, and considering this, incentives provided at the 1st and 4th visits were excessive. Further technical details, including statistical assumptions required for the encoding framework and further examples were included in the Appendix 3. A Python implementation including example code to calculate off-policy evaluation estimators for any user-specified CM parameter was provided at our GitHub repository: https://github.com/kyg0910/Optimizing-Contingency-Management-with-Reinforcement-Learning.

### Appendix 2. Details on the Search Grid

For the prize probabilities, we considered points on convex combinations between prize probabilities for usual care (UC) and for CM schema used in CTN-0007,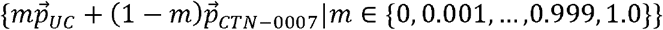 where 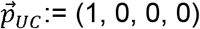 was the vector for UC, i.e., probability of 1 for no prize, and 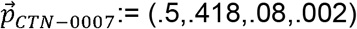 was for the CM schema used in CTN-0007. The larger *m* indicates that allocating higher probabilities on the no prize. Figure 2(a) visualized example values of prize probabilities.

The choice of values in logarithmic and logistic rules, 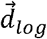 and 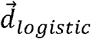, was to approximate 3.2ln *L* + 1 and 13/(1 + exp,(_0.69(*L* _6.5))), respectively, where *L* denoted the longest duration of substance abstinence in the week. The logarithmic curve passed the initial point of the linear escalation, (1, 1), and the logistic curve was rotationally symmetric with respect to (6.5, 6.5), the midpoint of the linear escalation curve, to have ticket numbers comparable to the linear rule in the middle stage and to move tickets in the early stage to the late stage. Figure 3(b) visualized escalation rules.

### Appendix 3. Data Preprocessing

*Data Acquisition*: The dataset for this study was obtained from CTN data repository. Three primary data files were used: Intake assessment questionnaire (corresponding to “qs.csv” data file), inclusion/exclusion criteria (“sc.csv”), and study termination (“suppds.csv”) forms. *Inclusion and Exclusion Criteria*: Our study adhered to the inclusion and exclusion criteria outlined in the CTN-0007 study protocol. We considered patients who had completed a minimum of four weeks of methadone treatment. Specifically, we included patients who met one of two conditions: (i) Those who had tested positive for stimulants in their urine within two weeks of the study entroy date and (ii) those who directly cam to the program from a controlled environment and had a stimulant positive urine in the two weeks before being enrolled in that environment. Participants who failed to provide informed consent during the simple consent quiz or had gambling problems were excluded. Additionally, we excluded participants with other issues, including those who ran in a methadone clinic before the research assistant’s arrival and cases where there was a lack of documentation to ascertain whether participants had completed the program. The final sample size of control and incentive groups were 188 and 195, respectively, and we used the incentive group in the analysis.

*Data Encoding*: Data encoding was performed as follows. For substance use histories, we used urine drug screens for amphetamine, cocaine, methamphetamine, and opioids. Alcohol status was determined based on breathalyzer readings, with readings smaller than or equal to 0.0009 considered negative, in accordance with the CTN-0007 study protocol. We encoded the primary test result as negative when all primary substances (alcohol, amphetamine, cocaine, and methamphetimne) tested negative. The secondary test result was considered negative when the opioid result was negative. All the other cases, including missing data, were encoded as positive.

For the lottery results, we recorded the number of draws for each of the four prize categories and the number of incentives provided.

## Footnotes

1 For each pair of estimators from two groups, say *A*_*j*_ from the first group and *B*_*k*_from the second, we computed the difference *A*_*j*_ −*B*_*k*_. Since there were 1,000 bootstrap importance sampling estimators in each group, this resulted in 1,000,000 computed differences. The p-value was the proportion of positive differences.

