## Supplementary material for "Optimizing Contingency Management with Reinforcement Learning": tables

**Table 1: Summary of trial settings and cohort characteristics of CTN-0007. For age, we presented means with standard deviations.**

| Aspect | Description |
| --- | --- |
| Sample size | Usual care ($n$=198), Usual care + CM ($n$=190) |
| Age | 42.0 (8.6) |
| Proportion of Females | 44.1% |
| Primary target substances | Alcohol, Amphetamine, Cocaine, Methamphetamine |
| Secondary target substances | Opioids |
| Main treatment outcomes | Retention, counseling attendance, the total number of stimulant- and alcohol-free samples provided, percentage of stimulant- and alcohol-negative samples provided, and the longest duration of abstinence. |

| **Table 2: Comparison of treatment outcomes and cost-effectiveness from various CM schemas. a: Upper bound of 95% bootstrap CIs for the CM in CTN-0007; b: Annual limit set by Washington State.** | | | | | | | | | | | | |
| --- | --- | --- | --- | --- | --- | --- | --- | --- | --- | --- | --- | --- |
| CM schemas | Parameter | | | | | |  | Outcome | | | | |
|  | prize probabilities | | | |  | escalation rule |  | incremental cost per participant | |  | the number of primary substance-free samples | |
|  | $0 | $1 | $20 | $80 |  |  |  | mean (SD) | 95% CI |  | mean (SD) | 95% CI |
| CM used in CTN-0007 | 50.00% | 41.80% | 8.00% | 0.20% |  | linear |  | $118 ($15) | [$92, $149] |  | 8.61 (0.68) | [7.36, 9.98] |
| Optimal CM when the budget is $149^a^ | 55.40% | 37.28% | 7.14% | 0.18% |  | logistic |  | $122 ($13) | [$98, $149] |  | 11.38 (0.93) | [9.59, 13.23] |
| Optimal CM when the budget is $100^b^ | 66.10% | 28.34% | 5.42% | 0.14% |  | logistic |  | $80 ($9) | [$63, $100] |  | 9.99 (0.83) | [8.44, 11.68] |

**Table 3: An example of data expressed with our reinforcement learning framework. The N and P indicate negative and positive, respectively. The reward is whether the test result at the next visit is negative. For the budget, we used the annual limit by Washington State, $100.**

| Visit time ($t$) | Test results up to and including the visit (state; $S_{t}$) | | Incentives (action; $A_{t}$) | Primary substance test result at the next visit - reward ($R_{t}$) | Remaining budget among $100 |
| --- | --- | --- | --- | --- | --- |
|  | Primary substances | Secondary substances |  |  |  |
| 1 | N | N | $20 | N - 1 | $80 |
| 2 | N, N | N, N | $2 | N - 1 | $78 |
| 3 | N, N, N | N, N, P | $1 | N - 1 | $77 |
| 4 | N, N, N, N | N, N, P, N | $21 | N - 1 | $56 |
| 5 | N, N, N, N, N | N, N, P, N, N | $0 | P - 0 | $56 |
| 6 | N, N, N, N, N, P | N, N, P, N, N, P | $0 | P - 0 | $56 |
