## Supplementary material for "Optimizing Contingency Management with Reinforcement Learning": figures

**Figure 1.** Illustration to describe our reinforcement learning framework for CM. It shows repeated motivational incentive allocations involving the CM schema (corresponding to "policy" in the Reinforcement Learning (54) denoted by $\pi_{\theta}^{t}$ where $\theta$ is the CM parameter) and patients during the treatment program. This repeated process reinforces patients' substance abstinence behavior using incentives and is fully characterized by the sequence of decision rules $(\pi_{\theta}^{1},\pi_{\theta}^{2},\ldots, \pi_{\theta}^{T})$ (corresponding to "dynamic treatment regimen" in precision medicine (51)) where $T$ is the number of total visits. **Action** (upper orange arrow): At the $t$-th visit, the CM schema allocates incentives ("action" denoted by $A_{t}$) based on substance use histories ("state" denoted by $S_{t}$). The incentives are expected to reinforce substance abstinence. **Observation** (lower green arrow): At the next stage, urine drug screens and breathalyzer samples are observed to check whether the participants abstained from the substances ("reward" denoted by $R_{t}$), and the substance use histories are updated to $S_{t+1}$, which the CM schema will use to allocate incentives at the next stage.

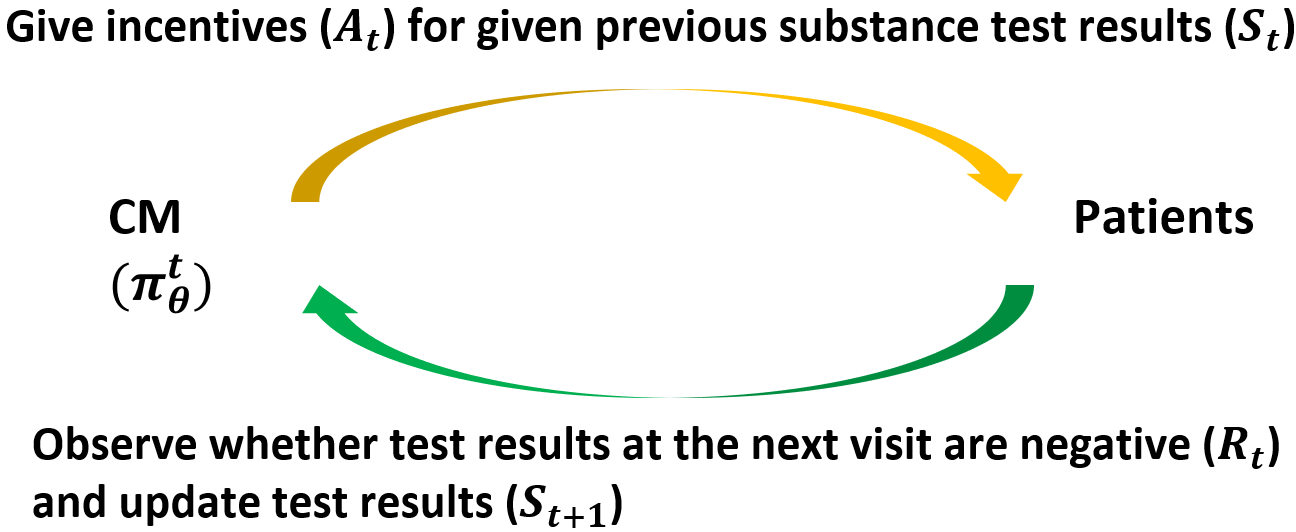

**Figure 2.** Illustration of CM parameters with the search grid and comparisons between costs and the number of primary substance-free samples from various CM schemas, including existing and optimal ones under the budget used in CTN-0007. **Panel (a)** displays the search grid for CM parameters, four-dimensional vectors for prize probabilities $\vec{p}$ in $x$-axis and twelve-dimensional vectors for escalation rules for the number of draws $\vec{d}$ in $y$-axis. The top, middle, and bottom horizontal lines show parameter values whose $\vec{p}$ parts are convex combinations between prize probabilities of usual care (UC) $\vec{p}_{UC}$and of the CM schema in CTN-0007 $\vec{p}_{CTN-0007}$ and $\vec{d}$ parts are fixed as vectors for linear rule $\vec{d}_{linear}$, logarithmic rule $\vec{d}_{log}$, and logistic rule $\vec{d}_{logistic}$, respectively. The CM parameter for the schema in CTN-0007, $(\vec{p}_{CTN-0007}, \vec{d}_{linear})$, is marked as a blue circle, and the optimal parameter under the budget used in CTN-0007, $\hat{\theta}^{*}=(0.108\vec{p}_{UC}+0.892\vec{p}_{CTN-0007}, \vec{d}_{logistic})$, is marked as a purple star. **Panel (b)** compares importance sampling estimators from CM schemes with diverse $\vec{p}$ while fixing $\vec{d}$ as $\vec{d}_{linear}$. Ellipses show 95% bootstrap confidence regions, using the means and covariances from 1,000 bootstrap estimators for cost and the number of primary substance-free samples, and bars at the top and right boundaries show boxplots of cost and the number of primary substances, respectively. The red dashed vertical line displays the upper bound of 95% bootstrap confidence intervals of cost from the CM schema used in CTN-0007, which we defined as the spent budget. **Panel (c)** compares CM schemes with diverse $\vec{d}$ while fixing $\vec{p}$ as $\vec{p}_{UC}$. Ellipses, bars, and the vertical line have the same meaning as in Panel (b). **Panel (d)** compares the existing CM parameter with the optimal one within the budget used in CTN-0007. Ellipses, bars, and the vertical line have the same meaning as in Panel (b).

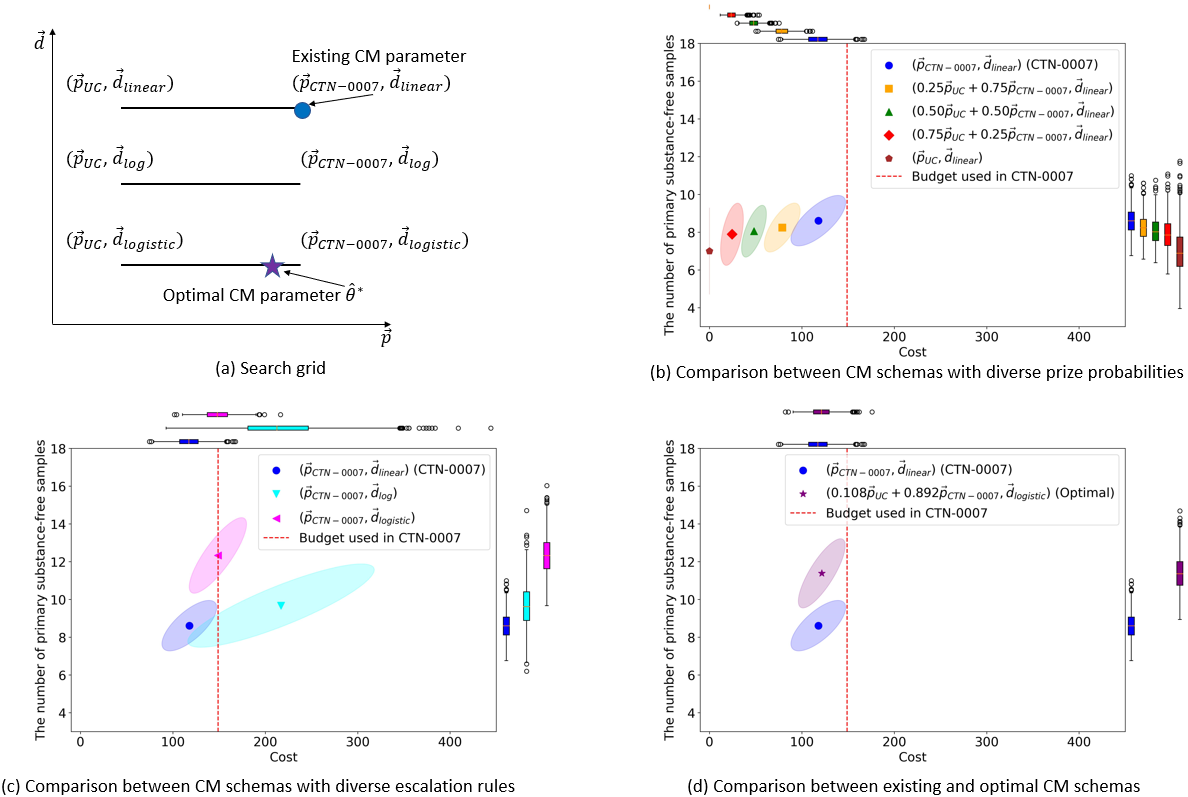

**Figure 3.** Visualization of two parameters, prize probabilities and escalation rules for the number of draws. **Panel (a)** displays five values of the four-dimensional prize probability vector $\vec{p}$. Each bar displays the proportions of probabilities of getting each prize from convex combinations between the vector for usual care (UC), $\vec{p}_{UC}:=(1,0,0,0)$, and for the CM schema in CTN-0007, $\vec{p}_{CTN-0007}:=(0.5, 0.418, 0.08, 0.002)$. The $x$-axis displays coefficients of convex combinations. **Panel (b)** displays three values of the twelve-dimensional number of draws vector $\vec{d}$. The $x$- and $y$-axis show the longest duration of (primary) substance abstinence in weeks and the corresponding number of draws, respectively. The blue line with circle points, orange line with square points, and green line with triangle points shows the number of draws for the linear rule $\vec{d}_{linear}:=(1, 2, \ldots, 11,12)$, logarithmic rule $\vec{d}_{log}:=\left( 1,4,\ldots,8,8 \right)$, and logistic rule $\vec{d}_{logistic}:=(1,1,\ldots,12,12)$, respectively.

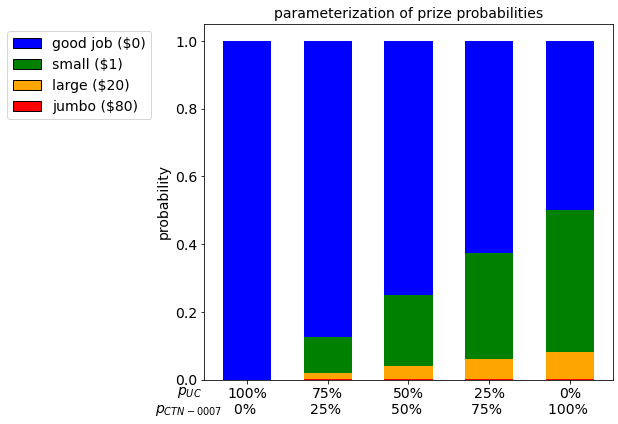

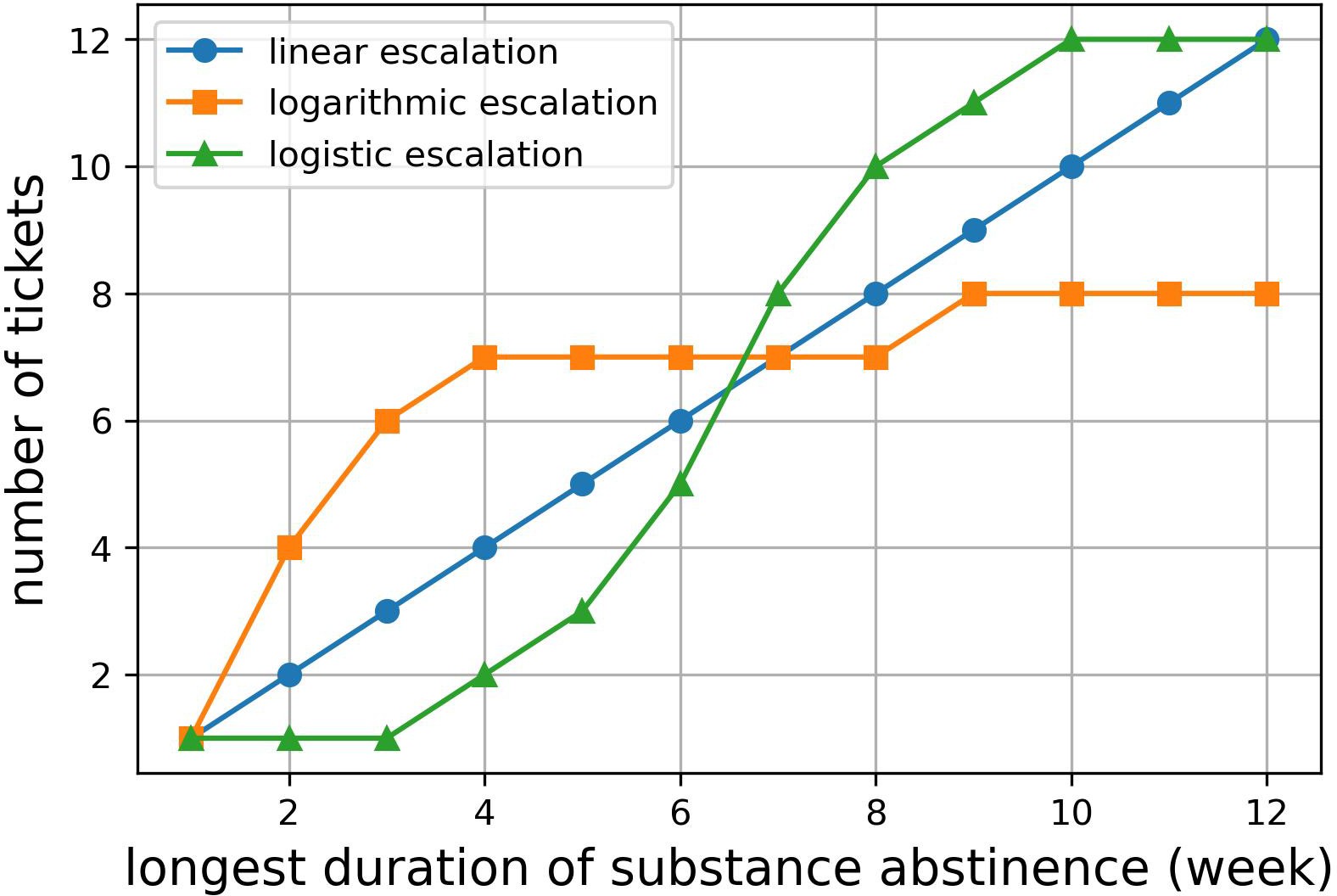

**(a)** Visualization of prize probabilities	**(b)** Visualization of escalation rules
